# SARS-CoV-2 IgG Antibody Responses in New York City

**DOI:** 10.1101/2020.05.23.20111427

**Authors:** Josh Reifer, Nosson Hayum, Benzion Heszkel, Ikey Klagsbald, Vincent A. Streva

## Abstract

Severe Acute Respiratory Syndrome Coronavirus 2 (SARS-CoV-2) is a strain of coronavirus that causes coronavirus disease 2019 (Covid-19) and has been declared a global pandemic by the World Health Organization. Total cases of SARS-CoV-2 worldwide exceed 4.8 million, with over 320,000 deaths recorded. Little is known about the body’s immune response to SARS-CoV-2 infection. In this paper, we describe SARS-CoV-2 IgG antibody responses in 28,523 patients from the New York City metropolitan area and report a SARS-CoV-2 IgG positivity rate of 44%, indicating the widespread nature of the pandemic in the city and state of New York. Additionally, for a subset of patients, we report on the correlation between SARS-CoV-2 patient symptom severity and level of SARS-CoV-2 IgG antibody found in the patient sample.

## INTRODUCTION

In December 2019, a novel strain of coronavirus was identified in a cluster of pneumonia patients in Huabei Province China in the city of Wuhan (1). This novel coronavirus strain [since named Severe Acute Respiratory Syndrome Coronavirus 2 (SARS-CoV-2)] is a member of the Betacoronavirus family and has ~80% sequence similarity to SARS-CoV-1, which is no longer circulating in humans (2). On March 11, 2020, the World Health Organization declared SARS-CoV-2 a global pandemic (3). Since its first description, SARS-CoV-2 has spread from China, and as of today (May 19, 2020) has spread to 215 countries and territories across the globe and is responsible for 4.81 million confirmed infections and over 320,000 deaths, including over 90,000 deaths in the United States (4,5).

SARS-CoV-2 spreads mainly though contact with infectious respiratory droplets generated when an infected person coughs or sneezes, or through contact with saliva or nasal discharge (6). Symptoms of infection by SARS-CoV-2 can range from mild (cough, fever, sore throat) to severe (shortness of breath, difficulty breathing, pneumonia) and more severe illness seems to affect the elderly or immunocompromised (7).

Little is known about the antibody response to SARS-CoV-2 infection. Antibodies against SARS-CoV-2 have been reported to be detected as early as a few days, to as many as three weeks after onset of symptoms (8,9), with the median time from symptom onset to detectable levels of IgG reported as six days (10). The presence of SARS-CoV-2 IgG antibodies generally are indicative of current or previous infection by SARS-CoV-2, and are thought to confer some degree of immunity (11), however the extent and duration of immunity conferred by SARS-CoV-2 IgG antibodies remains unknown (10,12).

A number of *in vitro* diagnostics have been developed for the detection of antibodies against SARS-CoV-2, and many fraudulent or ineffective tests have been released into the market (13).

Recently, a number of SARS-CoV-2 antibody tests have received Food and Drug Administration (FDA) Emergency Use Authorization (EUA) (14). These tests detect either individual SARS-CoV-2 specific IgG antibodies or total antibodies against SARS-CoV-2. All FDA EUA SARS-CoV-2 antibody tests target antibodies against either the viral nucleocapsid protein or spike glycoproteins, which are the two most abundant immunogenic components produced by the virus during infection (15). Much remains to be seen whether nucleocapsid or spike glycoprotein is a better target antigen when developing a SARS-CoV-2 antibody assay, however there is reason to suspect that in the future, spike glycoprotein will be preferred as a target, as the viral nucleocapsid is highly homologous among all strains of *Coronaviridae* (16) and the spike glycoprotein has been shown to be the target of neutralizing antibodies (17-19).

Little is known about how the body’s antibody response varies with severity of SARS-CoV-2 symptoms, however it is expected that more severe SARS-CoV-2 infections will result in more robust production of SARS-CoV-2 IgG antibodies. In this study, we report on the semi-quantitative SARS-CoV-2 IgG antibody levels in a population of patients scored for severity of illness and show a direct correlation between SARS-CoV-2 IgG antibody levels and severity of symptoms.

## METHODS

### Patient Populations

Patients were seen at primary care providers and urgent care facilities throughout New York city and the surrounding suburbs (Westchester, Rockland, Orange, Nassau, and Suffolk counties). The majority of patients (74%) were residents of the five boroughs (counties) that make up New York City (Kings (Brooklyn), Queens, New York (Manhattan), Bronx, and Richmond (Staten Island)). A subset of 240 patients were used to correlate symptom severity to antibody response. This subset of patients were seen for SARS-CoV-2 IgG testing at a single primary care provider in the Boro Park neighborhood of Brooklyn over a one month period from May 5, 2020 through June 5, 2020. Patients were tested for SARS-CoV-2 IgG between eight and twelve weeks after known or probable exposure to SARS-CoV-2.

### Clinical Patient Scoring (Symptom Index)

Patients were seen at an urgent care facility in Brooklyn, New York in May and early June 2020. As part of their screening, healthcare providers at this facility were asked to rate the severity of patients’ symptoms on a scale of 0 to 4 (Table 1) to generate a ‘Symptom Severity Index’ (SSI) to be used as a marker of severity of disease. All SSI were generated before the specimen was sent to the laboratory for testing, so the SSI result is generated independently of the SARS-CoV-2 IgG result.

**Table 1.** Description of Symptom Severity Index (SSI). Patients were assigned an SSI based on the criteria in Table 1. The SSI was assigned at the time of the patient’s office visit by the healthcare provider before laboratory testing for SARS-CoV-2 IgG.

| SSI | Description |
| --- | --- |
| 0 | Asymptomatic patient with no known exposure to SARS-CoV-2 positive patient(s) |
| 1 | Asymptomatic patient with known exposure to SARS-CoV-2 positive patient(s) |
| 2 | Mildly symptomatic patient (fever, cough, sore throat) with no pneumonia or drop in oxygen saturation levels |
| 3 | Symptomatic patient including unconfirmed pneumonia; drop in oxygen saturation as low as 92% |
| 4 | Severely symptomatic patient including x-ray confirmed pneumonia; oxygen saturation below 92% |

### Laboratory Testing for SARS-CoV-2 IgG

Semi-quantitative SARS-CoV-2 IgG testing was performed at Sherman Abrams Laboratory in Brooklyn, NY on serum specimens using the DiaSorin LIAISON SARS-CoV-2 S1/S2 IgG Antibody Assay on the LIAISON XL platform. This assay is an FDA EUA diagnostic test for the qualitative detection of SARS-CoV-2 IgG antibodies but produces a numerical index value (in arbitrary units per milliliter) which is indicative of the intensity of the reaction, and therefore a surrogate for SARS-CoV-2 IgG antibody levels.

### Data Analysis

Statistical analyses were performed using R. Plots of IgG index and age versus SSI were generated in R, using ggplot, with a data smoothing line applied. Linear regression analysis was performed using the lm() function native to R.

## RESULTS

During the study period, we generated a Symptom Severity Index (SSI) and semi-quantitative SARS-CoV-2 IgG level on 240 patients. Patient symptoms were scored by the healthcare provider at the time of office visit from 0-4 using criteria predetermined before the start of the study (Table 1). Of the 240 patients assigned an SSI, 36% were scored as zeroes, 35% were scored as ones, 21% were scored as twos, 5% were scored as threes, and 3% were scored as fours (Table 2).

**Table 2.** Number of Patients in each SSI group. Two hundred and forty patients were assigned an SSI at the time of clinic visit. Of these, 36% were assigned an SSI of 0, 35% were assigned an SSI of 1, 21% were assigned an SSI of 2, 5% were assigned an SSI of 3, and 3% were assigned an SSI of 4.

| SSI | Number | Percentage |
| --- | --- | --- |
| 0 | 86 | 36 |
| 1 | 85 | 35 |
| 2 | 51 | 21 |
| 3 | 12 | 5 |
| 4 | 6 | 3 |
| TOTAL | 240 | 100 |

The distribution of male and female patients in each of the SSI bins is shown in Table 3. There was no significant difference in gender distribution between patients with SSIs from zero to two, however patients with an SSI of three or four were disproportionately male. Whether this difference is due to increased disease severity among male patients (20) or the relatively small number of patients with an SSI of three or greater remains to be seen. Additionally, the median age of each SSI group was also determined (Table 4). Generally, patients with a lower SSI were younger than those with a higher SSI (Figure 1A), possibly indicative of more severe disease in older patients. This difference was true for both male and female patients (Figure 1B) and was statistically significant (p-value < 0.01) using a linear regression model.

**Table 3.** SSI and Patient Sex. Patient sex for each of the five SSI groups is shown. Patients in SSI groups 0 through 3 were roughly evenly split by patient sex, which those in SSI groups 3 and 4 tended to skew male.

| SSI | Patient Sex | Number | Percentage |
| --- | --- | --- | --- |
| 0 | Male | 44 | 51 |
|  | Female | 42 | 49 |
| 1 | Male | 44 | 52 |
|  | Female | 41 | 48 |
| 2 | Male | 26 | 51 |
|  | Female | 25 | 49 |
| 3 | Male | 7 | 58 |
|  | Female | 5 | 42 |
| 4 | Male | 5 | 83 |
|  | Female | 1 | 17 |

**Table 4.** Median Patient Age for each SSI group. The median patient age tends to increase with SSI. The median age for each SSI is shown as well as the median age by patient sex for each SSI.

| SSI | Patient Sex | Median Age (years) |
| --- | --- | --- |
| 0 | Total | 29 |
|  | Male | 29 |
|  | Female | 29 |
| 1 | Total | 35 |
|  | Male | 35 |
|  | Female | 35 |
| 2 | Total | 45 |
|  | Male | 45 |
|  | Female | 42 |
| 3 | Total | 48 |
|  | Male | 42 |
|  | Female | 56 |
| 4 | Total | 46 |
|  | Male | 45 |
|  | Female | 73 |

**Figure 1.**
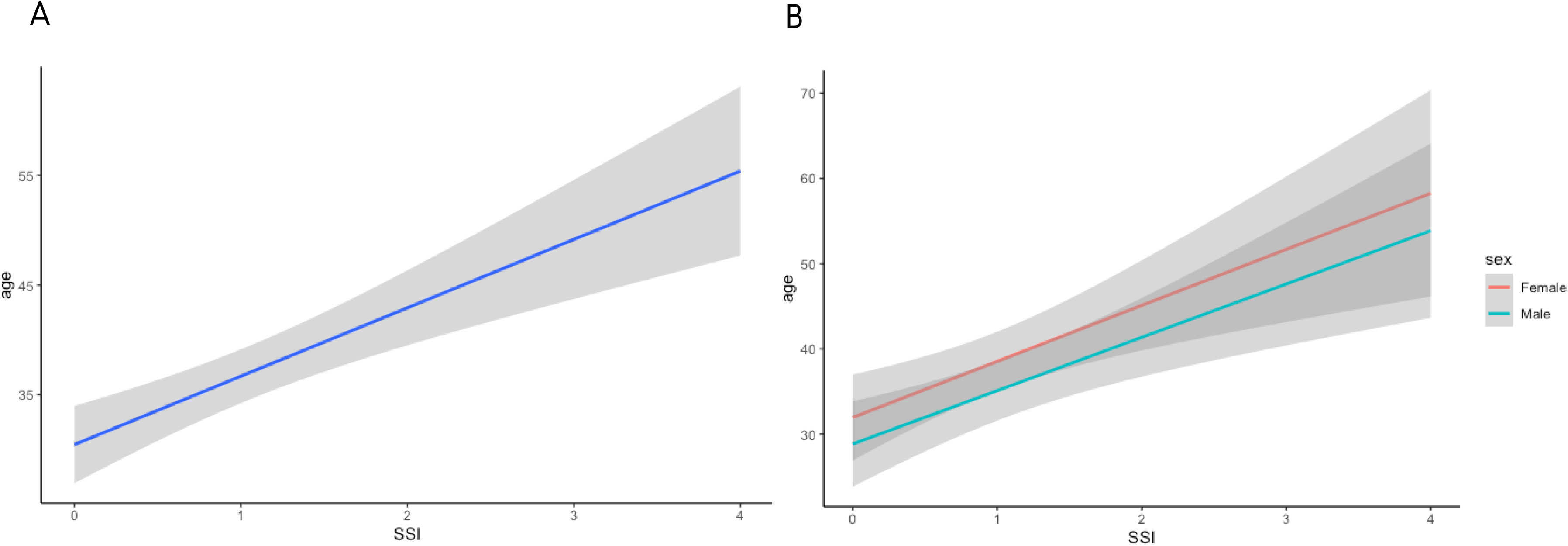
Age versus Symptom Severity Index. (A) Patient age is plotted versus symptom severity index (SSI) for all 240 patients given an SSI. Blue line represents the regression line and gray shading represents the 95% confidence interval of the regression. The correlation between age and SSI is statistically significant at p < 0.01 (B) Patient age is plotted versus symptom severity index (SSI) for all 240 patients given an SSI and broken down by patient sex. The linear regression line for males (blue line) and females (red line) along with 95% confidence intervals of the regressions (gray shading) are shown. The correlation between age and SSI is statistically significant for both male and female patients at p < 0.01

We next sought to evaluate whether semi-quantitative SARS-CoV-2 IgG antibody levels were correlated to severity of symptoms as measured by the SSI. Levels of SARS-CoV-2 IgG antibody are plotted against SSI in Figure 2A. A linear regression analysis of the data indicates that SARS-CoV-2 IgG antibody levels are positively correlated with SSI (p-value < 0.01). This trend is true regardless of whether the patient was male or female (Figure 2B). These data imply that more severe SARS-CoV-2 infections are more likely than less severe infections to generate sufficient IgG antibodies to confer protective immunity.

**Figure 2.**
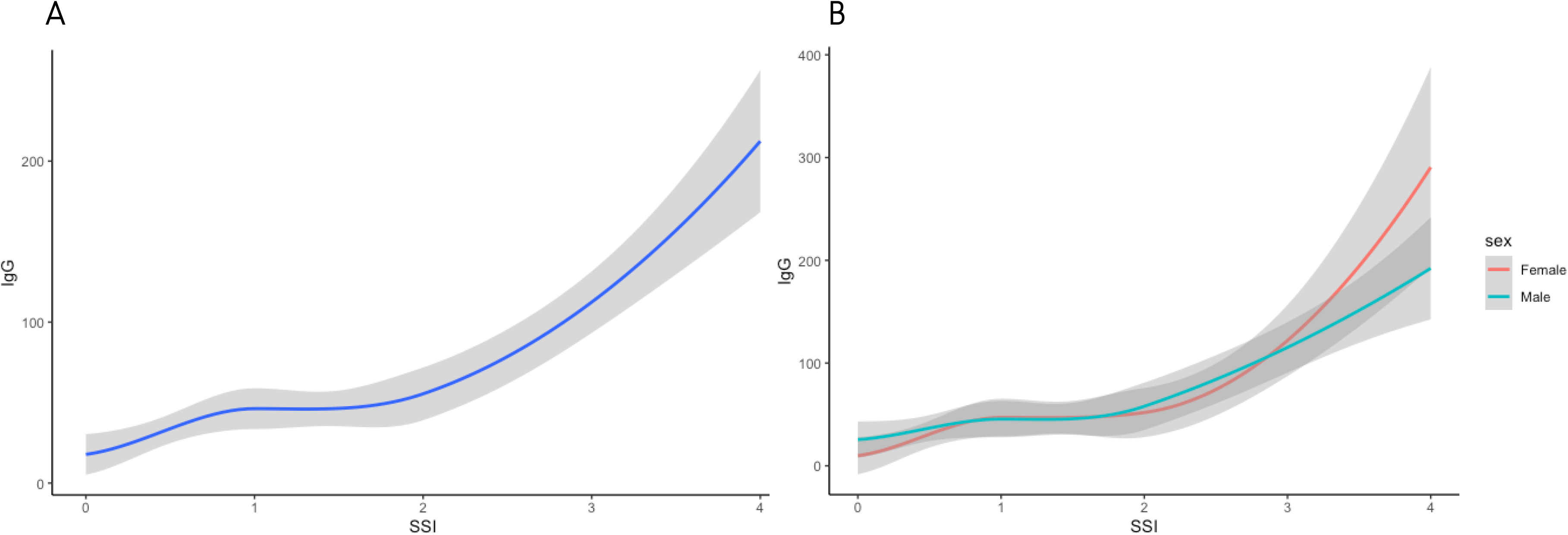
SARS-CoV-2 IgG Index Level versus Symptom Severity Index. (A) SARS-CoV-2 IgG index level (AU/mL) is plotted versus symptom severity index (SSI) for all 240 patients given an SSI. Blue line represents the regression line and gray shading represents the 95% confidence interval of the regression. The correlation between SARS-CoV-2 IgG level and SSI is statistically significant at p < 0.01 (B) SARS-CoV-2 IgG index level (AU/mL) is plotted versus symptom severity index (SSI) for all 240 patients given an SSI and broken down by patient sex. The linear regression line for males (blue line) and females (red line) along with 95% confidence intervals of the regressions (gray shading) are shown. The correlation between SARS-CoV-2 IgG level and SSI is statistically significant for both male and female patients at p < 0.01

In addition, we tested 28,523 patient specimens for SARS-CoV-2 IgG antibody levels for whom we did not obtain a SSI. Of these, 12,424 were SARS-CoV-2 IgG positive indicating a 44% positivity rate in our patient population, which reflects the extent of the Covid-19 pandemic in New York City. Positivity rates in males were higher than in females (49% positive IgG in male patients, and 38% positive IgG in female patients). The lowest prevalence of SARS-CoV-2 IgG was in patients aged 86-90, with a positivity rate of 25% and pattients aged zero through five, with a positivity rate of 26% (note, there were no positive IgG results in patients 101-105 years old, but only three patients from this demographic were tested, so they are omitted from this analysis). Interestingly, the age groups with the highest prevalence of positive SARS-CoV-2 IgG are ages 11-15 (56% positive) and ages 16-20 (57% positive) (Figure 3).

**Figure 3.**
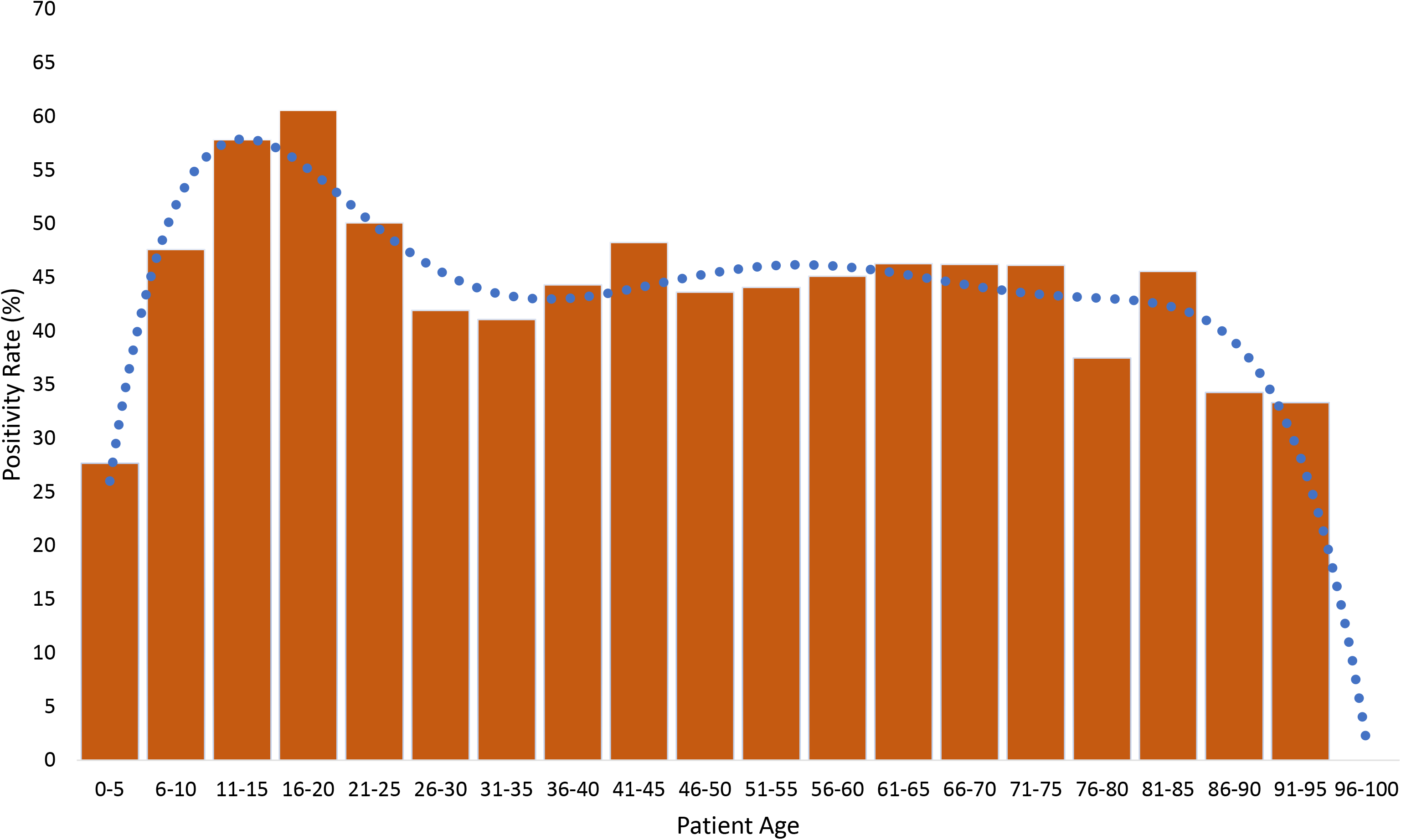
Patient SARS-CoV-2 IgG positivity rates. Histogram of SARS-CoV-2 IgG positivity rate by patient age for 28,523 patients from the New York City metropolitan area. Patients were binned into five year age groups and the positivity rate for each age group is plotted. SARS-CoV-2 IgG positivity rates ranged from 24.5% in patients aged 86-90 to 57.3% in patients aged 16 to 20 years. The blue dotted line represents a fitted regression line intended to show trends.

We also calculated the mean semi-quantitative IgG Index value by age for the 12,424 SARS-CoV-2 IgG positive patients tested. The mean index value across ages was 108.4 AU/mL, with the average index in males (112.7 AU/mL) being significantly higher than the average index in females (101.4 AU/mL) (p < 0.05). The mean SARS-CoV-2 IgG index value is lowest in young adults (ages 16-40) and is highest in children (ages zero to ten) and older adults (ages 76 and above) (Figure 4A and 4B). This observation is true for both males and females (Figure 4C and 4D).

**Figure 4.**
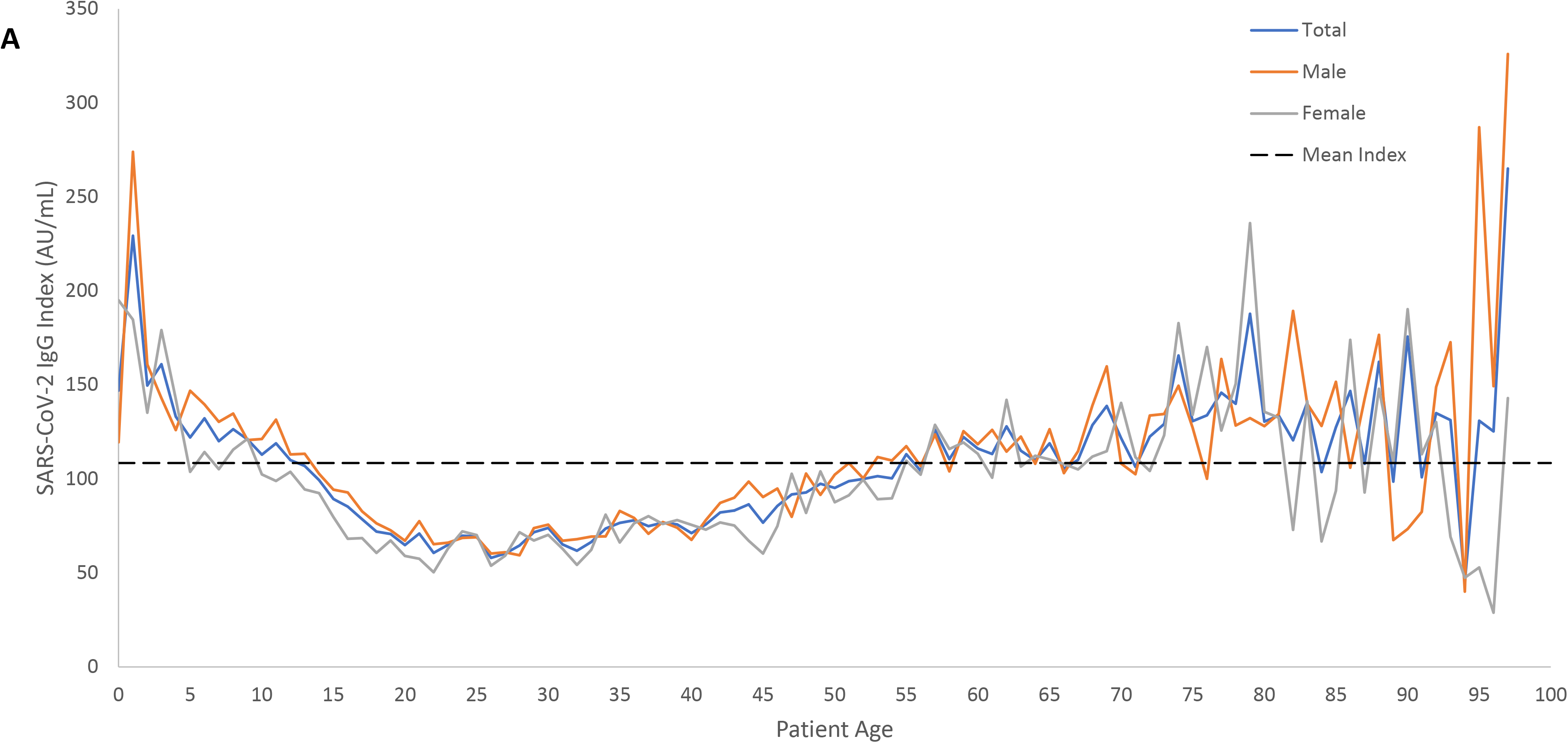
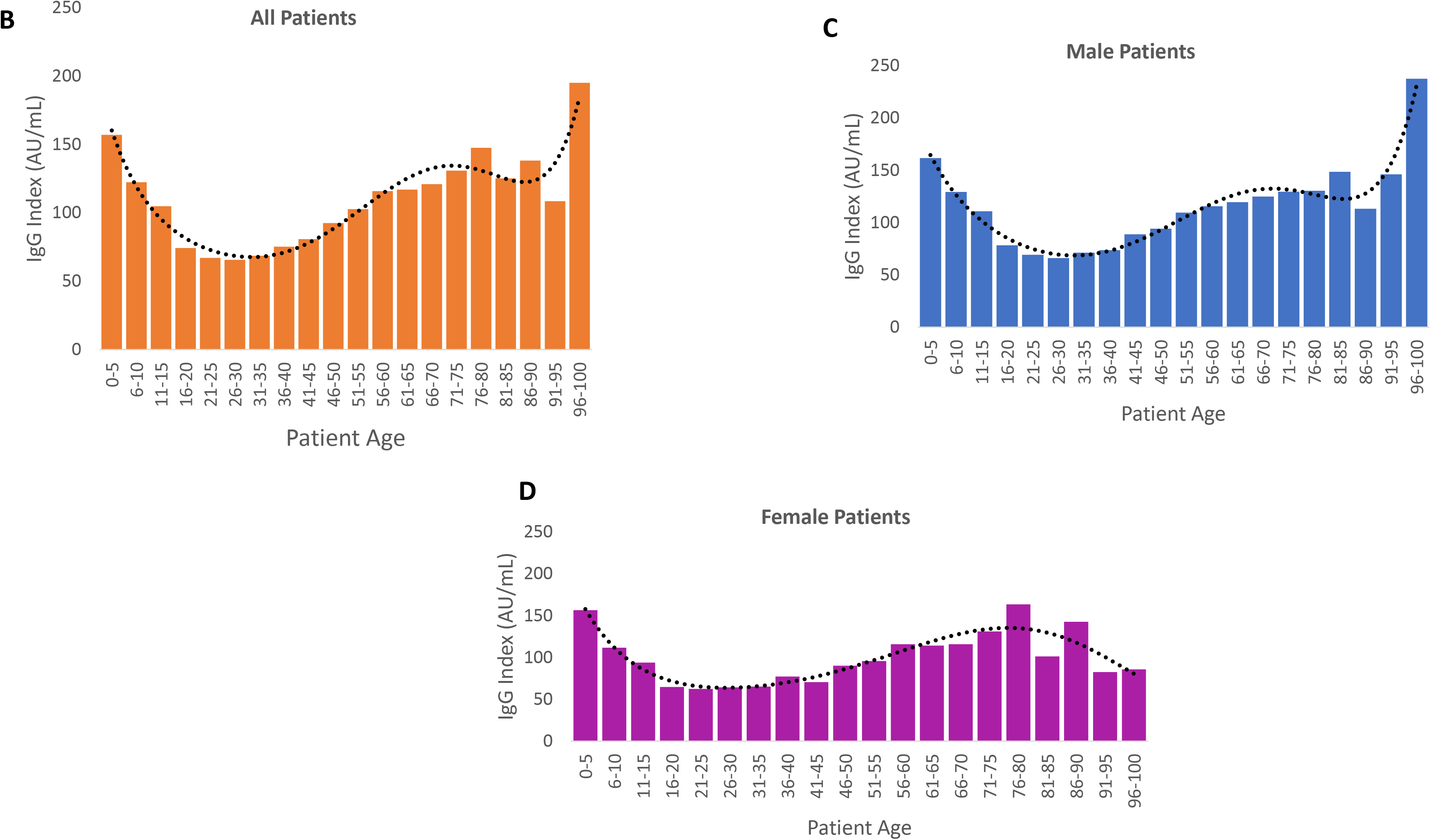
SARS-CoV-2 IgG Index Values by Age. (A) The mean SARS-CoV-2 IgG index values (AU/mL) of the 12,424 SARS-CoV-2 IgG positive patients in this study for each age are plotted versus patient age. The mean values for males (orange line), females (gray line), and both sexes (blue line) are plotted. The mean index value for all positive patients was 108.4 AU/mL (black dashed line). (B) The mean SARS-CoV-2 IgG index values (AU/mL) of the 12,424 SARS-CoV-2 IgG positive patients in this study were plotted versus patient age, with age binned into five year groups. The SARS-CoV-2 IgG index value is lowest in patients ages 16 to 40. The black dotted line is a regression line fitted to the data to show trends. (C) The mean SARS-CoV-2 IgG index values (AU/mL) of male SARS-CoV-2 IgG positive patients were plotted versus patient age, with age binned into five year groups. The SARS-CoV-2 IgG index value is lowest in patients ages 16 to 40. The black dotted line is a regression line fitted to the data to show trends. (D) The mean SARS-CoV-2 IgG index values (AU/mL) of female SARS-CoV-2 IgG positive patients were plotted versus patient age, with age binned into five year groups. The SARS-CoV-2 IgG index value is lowest in patients ages 16 to 35. The black dotted line is a regression line fitted to the data to show trends.

## DISCUSSION

This study is the first to correlate severity of SARS-CoV-2 symptoms with antibody response using an FDA EUA SARS-CoV-2 IgG assay. Additionally, it is one of the first to provide a snapshot of SARS-CoV-2 IgG antibody responses in a region of the country that was hardest hit by the virus, New York City through an analysis of over 28,000 patients presenting to primary care providers for SARS-CoV-2 IgG antibody testing.

We show SARS-CoV-2 IgG positivity rates in our patient population to be nearing 50%, which is indicative of the widespread nature of this pandemic in New York City. Further, our data show that the highest rates of prior infection are seen in patients aged 11-20 years, however it is formally possible that part of the explanation for this is a more robust immune response in this generally young, healthy age group.

Additionally, while the data are preliminary, our study shows that patients who experienced a more severe clinical course of SARS-CoV-2 infection are more likely to have higher serum levels of SARS-CoV-2 IgG antibody. Therefore, it follows, that these individuals are more likely to benefit from protective immunity and are less prone, at least in the short-term, to reinfection by SARS-CoV-2.

One potential limitation of our study, is the potential for waning immunity in the weeks after infection. The patient cohort used to correlate SSI to SARS-CoV-2 antibody levels were from a single tight-knit community in the Boro Park neighborhood of Brooklyn, and were epidemiologically determined to have had exposure or expected exposure to SARS-CoV-2 eight to twelve weeks prior to presenting for SARS-CoV-2 IgG testing. While we cannot entirely rule out alternative time-courses of infection in all patients, the clinical and epidemiological data gathered suggest all patients in the SSI cohort were infected by SARS-CoV-2 within approximately two weeks of one another. From the larger sample of 28,523 patients, we do not have access to sufficient epidemiological data to conclusively determine suspected dates of exposure to SARS-CoV-2. Future work will be needed to track SARS-CoV-2 IgG antibody levels over time.

The findings of our study have additional implications for blood banks seeking donors for convalescent plasma to treat Covid-19 patients. These facilities have traditionally screened donors for antibodies using qualitative antibody tests and have often relied on self-reporting by patients of their history and timing of symptoms to support a previous diagnosis of SARS-CoV-2. The availability of a semi-quantitative SARS-CoV-2 IgG assay with index values that correlate to severity of past infection could prove to be of significant help in selecting plasma donors.

Additional studies are underway to better understand the timing of the IgG antibody response to SARS-CoV-2 and how long those antibodies are maintained in circulation. Further, studies to correlate the semi-quantitative IgG Index value given by the DiaSorin LIAISON test to a direct measure of viral neutralization (eg, plaque reduction neutralization titer) are ongoing. Until this information is available, our study provides clinicians with information they need to gauge antibody responses between patients and correlate those antibody values to understand the patient’s clinical history.

## Data Availability

All data in the manuscript are available

## ACKNOWLEDGEMENTS

The authors thank Dr. Dakai Liu and Sanford Moos for helpful discussions and critical reading of the manuscript, and Verniece Hudson, Rachelle Neri, Joyce Zhen, and Edward Chu for generating clinical laboratory testing results.

## AUTHOR CONTRIBUTIONS

JR conceived the study, collected and analyzed data, and revised the manuscript; NH gathered clinical data and revised the manuscript; BH and IK analyzed data and revised the manuscript; VAS conceived the study, analyzed data, and wrote the manuscript.

